# Brain-Wide Mendelian Randomization Study of Anxiety Disorders and Symptoms

**DOI:** 10.1101/2023.09.12.23295448

**Authors:** Mihaela-Diana Zanoaga, Eleni Friligkou, Jun He, Gita A. Pathak, Dora Koller, Brenda Cabrera-Mendoza, Murray B. Stein, Renato Polimanti

**Affiliations:** Department of Psychiatry, Yale University School of Medicine, New Haven, CT, USA; VA Connecticut Healthcare System, West Haven, CT, USA; Department of Genetics, Microbiology, and Statistics, Faculty of Biology, University of Barcelona, Catalonia, Spain; Department of Psychiatry, University of California, San Diego, La Jolla, CA, USA; Herbert Wertheim School of Public Health, University of California, San Diego, La Jolla, CA, USA; VA San Diego Healthcare System, San Diego, CA, USA; Wu Tsai Institute, Yale University, New Haven, CT, USA

**Keywords:** Causal Inference, Genome-wide Association Studies, Brain Structure, Brain Function, Pleiotropy

## Abstract

**Background:** To gain insights into the role of brain structure and function on anxiety (ANX), we conducted a genetically informed investigation leveraging information from ANX genome-wide association studies available from UK Biobank (UKB; N=380,379), FinnGen Program (N=290,361), and Million Veteran Program (MVP; N=199,611) together with UKB genome-wide data (N=33,224) related to 3,935 brain imaging-derived phenotypes (IDP).

**Methods:** A genetic correlation analysis between ANX and brain IDPs was performed using linkage disequilibrium score regression. To investigate ANX–brain associations, a two-sample Mendelian randomization (MR) was performed considering multiple methods and sensitivity analyses. A subsequent multivariable MR (MVMR) was executed to distinguish between direct and indirect effects. Finally, a generalized linear model was used to explore the associations of brain IDPs with ANX symptoms.

**Results:** After false discovery rate correction (FDR q<0.05), we identified 41 brain IDPs genetically correlated with ANX without heterogeneity among the datasets investigated (i.e., UKB, FinnGen, and MVP). Six of these IDPs showed genetically inferred causal effects on ANX. In the subsequent MVMR analysis, reduced area of the right posterior middle-cingulate gyrus (rpMCG; beta=-0.09, P= 8.01×10^-4^) and reduced gray-matter volume of the right anterior superior temporal gyrus (raSTG; beta=-0.09, P=1.55×10^-3^) had direct effects on ANX. In the ANX symptom-level analysis, rpMCG was negatively associated with “tense sore oraching muscles during the worst period of anxiety” (beta=-0.13, P=8.26×10^-6^).

**Conclusions:** This study identified genetically inferred effects generalizable across large cohorts, contributing to understand how changes in brain structure and function can lead to ANX.

## INTRODUCTION

Anxiety disorders (ANX) are mental illnesses characterized by excessive and persistent fear, panic, and anxious feelings (1). ANX is a public health burden due to its association with disability, poor quality of life, impaired daily functioning, and comorbidity with negative mental and physical health outcomes (2). These factors are among those that has led to the US Preventive Services Task Force (USPSTF) to recommend routine screening for anxiety disorders (3).

Previous studies found that the hypothalamic-pituitary-adrenal (HPA) axis is involved in ANX pathophysiology (4), with some affected individuals presenting alterations in cortisol levels (5, 6). Furthermore, advances in neurobiology and molecular genetics also highlighted the role of various neurotransmitters (7), such as serotonin and gamma-aminobutyric acid (GABA) (8, 9), as well as altered glutamate signaling (10) or increased norepinephrine levels in certain brain regions (11). ANX neuroimaging research identified activation patterns in brain regions related to emotion monitoring and processing (12, 13). For example, individuals with ANX showed increased volume or hyperactivation in the amygdala that may be related to heightened fear responses (14, 15), and reduced volume and dysregulation of the prefrontal cortex, potentially contributing to difficulties in regulating emotional responses (16, 17). Dysfunction in the anterior cingulate cortex has been reported as associated with increased activation in response to threat-related stimuli (18, 19). Similarly, alterations in hippocampal volume, connectivity, and functions have been linked to with memory deficits and emotional issues (20, 21). While these investigations provided important information regarding ANX pathogenesis, their limited sample size permitted only to investigate specific a priori hypotheses. Additionally, because of their cross-sectional design, their results cannot be interpreted in the context of possible cause-effect relationships.

In recent years, genome-wide association studies (GWAS) generated an unprecedented amount of information regarding psychiatric disorders and traits, also including ANX-related phenotypes (22). In addition to identifying potential risk loci, genetic associations can be used as an anchor to infer possible causal relationships. In particular, we can apply the two-sample Mendelian randomization (MR) approach to test the causal effect of a risk factor (i.e., exposure) on an outcome of interest using information regarding genetic associations available from GWAS (23). To investigate the possible causal effects linking brain structure and function to ANX, we leveraged ANX GWAS from UK Biobank (UKB; N=380,379) (24), FinnGen Program (N=290,361) (25), and Million Veteran Program (MVP; N=199,611) (26) together with UKB genome-wide data (N=33,224) related to 3,935 brain imaging-derived phenotypes (IDP) derived from six magnetic-resonance imaging (MRI) modalities (27).

## METHODS

### Data Sources

GWAS data investigated in the present study were generated from UKB, FinnGen, and MVP cohorts (24, 25, 28).

UKB is a population-based study that collected phenotypic and genotypic information regarding over 500,000 participants (24, 29). In the present study, we used ANX genome-wide association statistics generated by the Pan-UKB analysis (details available at https://pan.ukbb.broadinstitute.org/docs/qc). Briefly, ANX was defined based on phecode 300 “anxiety disorders” including 10,449 cases and 369,930 controls. UKB genome-wide association analysis was conducted using regression models available in Hail (available at https://github.com/hail-is/hail) and including the top-20 within-ancestry principal components (PC), sex, age, age^2^, sex×age, and sex×age^2^ as covariates. From UKB, we also derived genome-wide association statistics regarding 3,935 brain IDPs assessed in up to 33,224 participants. Details regarding IDP GWAS analyses have been previously described (27). UKB protocols for brain imaging acquisition, data pre-processing, and analysis techniques (30, 31) generated brain IDPs from six MRI modalities: T1-weighted structural imaging (T1), T2-weighted fluid-attenuated inversion recovery imaging (T2 FLAIR), Diffusion-weighted imaging (dMRI), Susceptibility-weighted imaging (swMRI), Resting-state functional MRI (rfMRI) and Task-based functional MRI (tfMRI).

FinnGen is a public-private research collaboration that collecting digitized medical records from Finnish health registries and genotyping data from samples available from Finnish biobanks (25, 32). In the present study, we used FinnGen genome-wide association statistics available from Release 8. A detailed description is available at https://finngen.gitbook.io/documentation/v/r8/. Briefly, GWAS was performed using REGENIE (33) including age, sex, top-10 within-ancestry PCs, and genotyping batch as covariates. FinnGen ANX definition was based on the medical endpoint “anxiety disorders” (KRA_PSY_ANXIETY) that included 35,385 cases and 254,976 controls.

MVP is a biobank funded by the U.S. Department of Veterans Affairs (VA) to study the health of U.S. military veterans, with over 900,000 veterans enrolled. Data are available through electronic health records, questionnaires, and blood samples (28). MVP ANX GWAS was conducted with respect to the total score of the generalized anxiety disorder 2-item scale (GAD-2) assessed in 199,611 individuals (26).

To maximize the power of our investigation while also assessing possible heterogeneity across the ANX GWAS datasets (i.e., UKB, FinnGen, and MVP), the analyses described below were conducted in each cohort separately and the results were combined via meta-analysis. This approach permitted us to estimate the heterogeneity among the cohort-specific effects in each analysis. The meta-analyses were computed using metafor R package (details available at https://wviechtb.github.io/metafor/reference/metafor-package.html), using the common effect model (34).

### Heritability and Genetic Correlation

Heritability (h2) and genetic correlation (rg) of ANX and brain imaging phenotypes were calculated using linkage disequilibrium score regression (LDSC) (35). The European populations available from the 1000 Genomes Project Phase 3 (36) were used as the LD reference panel. The default quality control criteria were applied: INFO > 0.9, MAF > 0.01, and 0 < P ≤ 1. Additionally, strand-ambiguous and insertion/deletion variants were removed.

### Two-Sample Mendelian Randomization

A two-sample Mendelian Randomization (MR) analysis (37) was performed using the TwoSampleMR R package (38) to estimate the bidirectional relationship between ANX (assessed in MVP and FinnGen) and brain IDPs (assessed in UKB). However, when testing the relationship between ANX and IDPs within UKB, MRlap approach (39) was applied to account for the possible bias introduced by the sample overlap between exposure and outcome datasets. In both analyses, variants associated with the exposures of interest considering a p=10^-5^ were selected as genetic instruments. To ensure that the variants were independent of each other, we clumped them using 1000 Genomes Project Phase 3 reference panel for the European populations and considering a window of 10,000 kb and a LD r2 cutoff of 0.001.

While we considered the inverse variance weighted (IVW) method as the primary due to its high statistical power, we compared IVW estimates from those obtained from other MR methods to evaluate the effect of possible biases. Because our study used a relaxed inclusion threshold (p=1e-5) to define the genetic instruments, we applied MR-RAPS (robust adjusted profile score) to account for the possible weak-instrument bias (40). MR-PRESSO (Mendelian Randomization Pleiotropy RESidual Sum and Outlier) was used to assess potential bias introduced by horizontal pleiotropy (41). Specifically, MR-PRESSO global test allowed us to detect genetic instruments affected by horizontal pleiotropy. MR-PRESSO outlier test was used to correct IVW estimates for biases introduced by horizontal pleiotropy, and the MR-PRESSO distortion test to evaluate distortion in the causal estimates before and after outlier removal (41). MR-PRESSO empirical p-values were calculated with respect to a null distribution from 10,000 permutations.

To investigate the independent effect of multiple exposures on the same outcome, a multivariable MR (MVMR) (42) approach was applied using the MendelianRandomization R package. In the MVMR analysis, genetic variants were selected as being associated with at least one exposure at p=10^-5^ and then clumped considering the same criteria described above.

### Anxiety Symptom-Level Analysis

To follow up on the findings of the MR analyses, the brain IDPs associated with ANX in the MR analysis were investigated with respect to 28 anxiety symptoms (Supplemental Table 1) assessed in the UKB mental health questionnaire (43). A generalized linear model (GLM) was fitted considering ANX symptoms as the dependent variable and brain IDPs as the independent variable together with age and sex as covariates. In addition to performing a sex-combined analysis, we also performed a sex-stratified analysis to explore possible differences between females and males.

## RESULTS

### Heritability and Genetic Correlation

ANX genetic correlation among the three cohorts investigated ranged from rg=0.61±0.04 between MVP and FinnGen to rg=0.80±0.07 between FinnGen and UKB. Details regarding LDSC statistics of each ANX GWAS are available in Supplemental Table 2. The cross-cohort meta-analysis of ANX genetic correlation identified 41 brain IDPs after false discovery rate multiple testing correction (FDR q<0.05; Figure 1). The ANX genetic correlation estimates observed for these IDPs did not show evidence of heterogeneity (heterogeneity-p>0.05; Supplemental Table 3). Among brain IDPs surviving FDR multiple testing correction, the most statistically significant ANX genetic correlations were observed for the median T2star in left pallidum (IDP 1444; rg=-0.13; p=6.21×10^-6^), the area of the right precentral gyrus (IDP: 0862; rg=-0.13, p=6.50×10^-6^), rfMRI connectivity ICA-feature 2 (IDP: 3915; rg=-0.12, p=6.80×10^-6^), and the right Brodmann area 6 (IDP: 0802; rg=-0.13, p=8.31×10^-6^).

**Figure 1.**
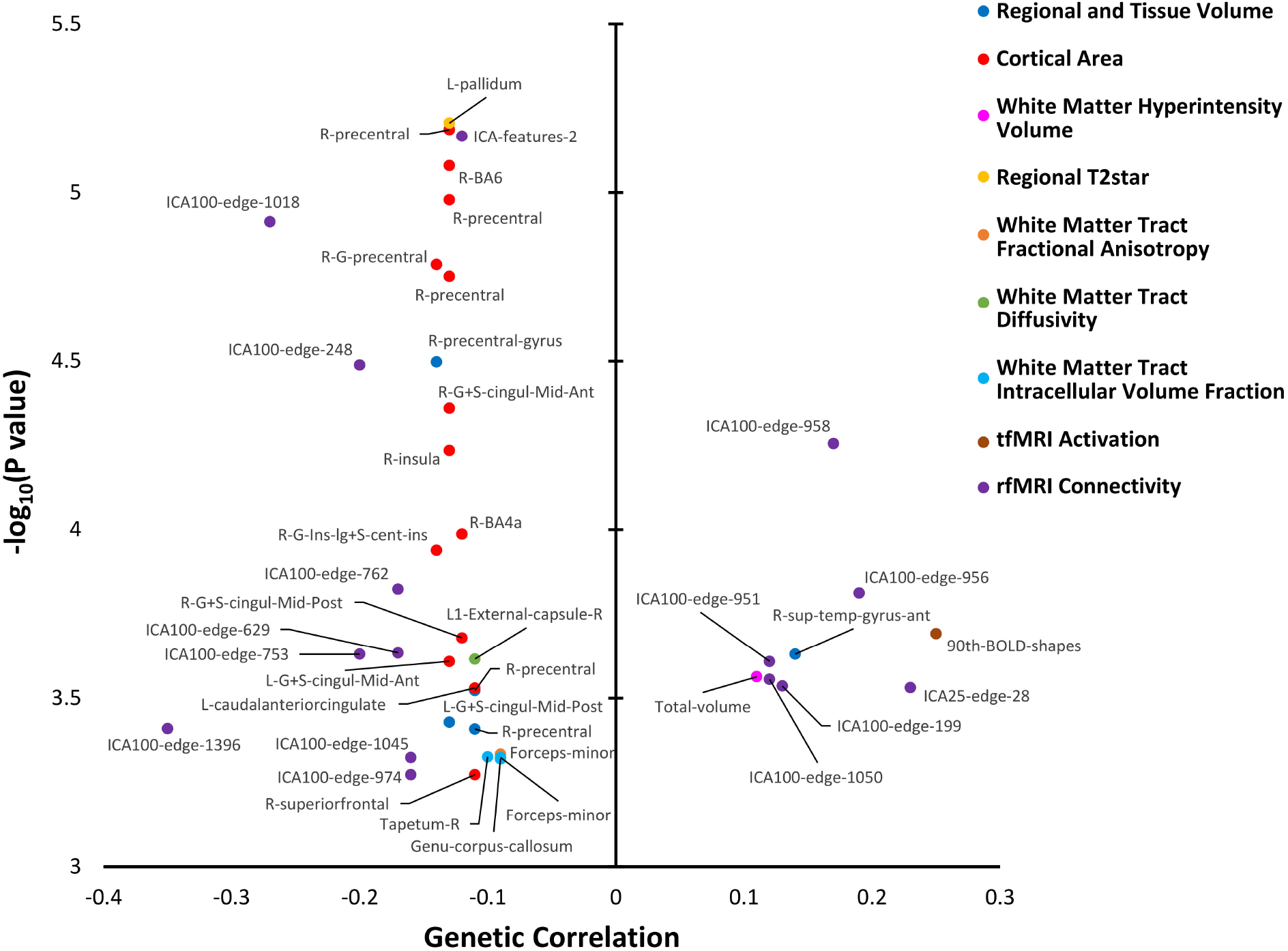
Genetic correlation between ANX and brain IDP surviving false discovery rate multiple testing correction in the cross-cohort meta-analysis (FDR q<0.05). Full description of the brain IDPs and their genetic correlation statistics are available in Supplemental Table 3.

### Two-Sample Mendelian Randomization

After multiple testing correction (FDR q<0.05), the bidirectional two-sample MR analysis identified eight brain IDPs causally associated with ANX (Supplemental Table 4), while ANX did not show any effect on the brain IDPs evaluated (p>0.05, Supplemental Table 5). In most cases, the effect of brain IDPs on ANX was negatively directed (i.e., smaller area, volume, or activity was associated with increased ANX; Figure 2). The only positive effect was related to the total volume of white matter hyperintensities (IDP 1437; beta=0.04, p=0.005). Among the effects identified, we observed statistically significant heterogeneity among ANX GWAS datasets for the area of the middle-anterior part of the right cingulate gyrus and sulcus (IDP 0952; heterogeneity-p=0.015) and the area of right superior-frontal gyrus (IDP 0709; heterogeneity=0.006). Because the effects related to these two IDPs may be driven by cohort-specific characteristics, we excluded them from further analyses. Conversely, for the remaining six IDPs, in addition to not observing heterogeneity across cohorts, the estimates of their effect on ANX were consistent among IVW, MR-RAPS, and MR-PRESSO approaches (Supplemental Figure 1). The six brain IDPs affecting ANX showed a certain degree of genetic correlation among them (Supplemental Table 6), ranging from rg=0.87±0.02 between the weighted-mean intra-cellular volume fraction in the forceps minor (IDP 1961) and the weighted-mean fractional anisotropy in the forceps minor (IDP 1511) to rg=-0.35±0.06 between the total volume of white matter hyperintensities (IDP 1437) and the weighted-mean intra-cellular volume fraction in the forceps minor (IDP 1961). Accordingly, we performed a MVMR analysis (Figure 3; Supplemental Table 7), observing independent effects for the grey-matter volume in the anterior division of the right superior temporal gyrus (IDP 0043; beta= -0.09, p=0.002) and the area of the middle-posterior part of the right cingulate gyrus and sulcus (IDP 0953; beta=-0.09 P-value=8.01×10^-4^).

**Figure 2.**
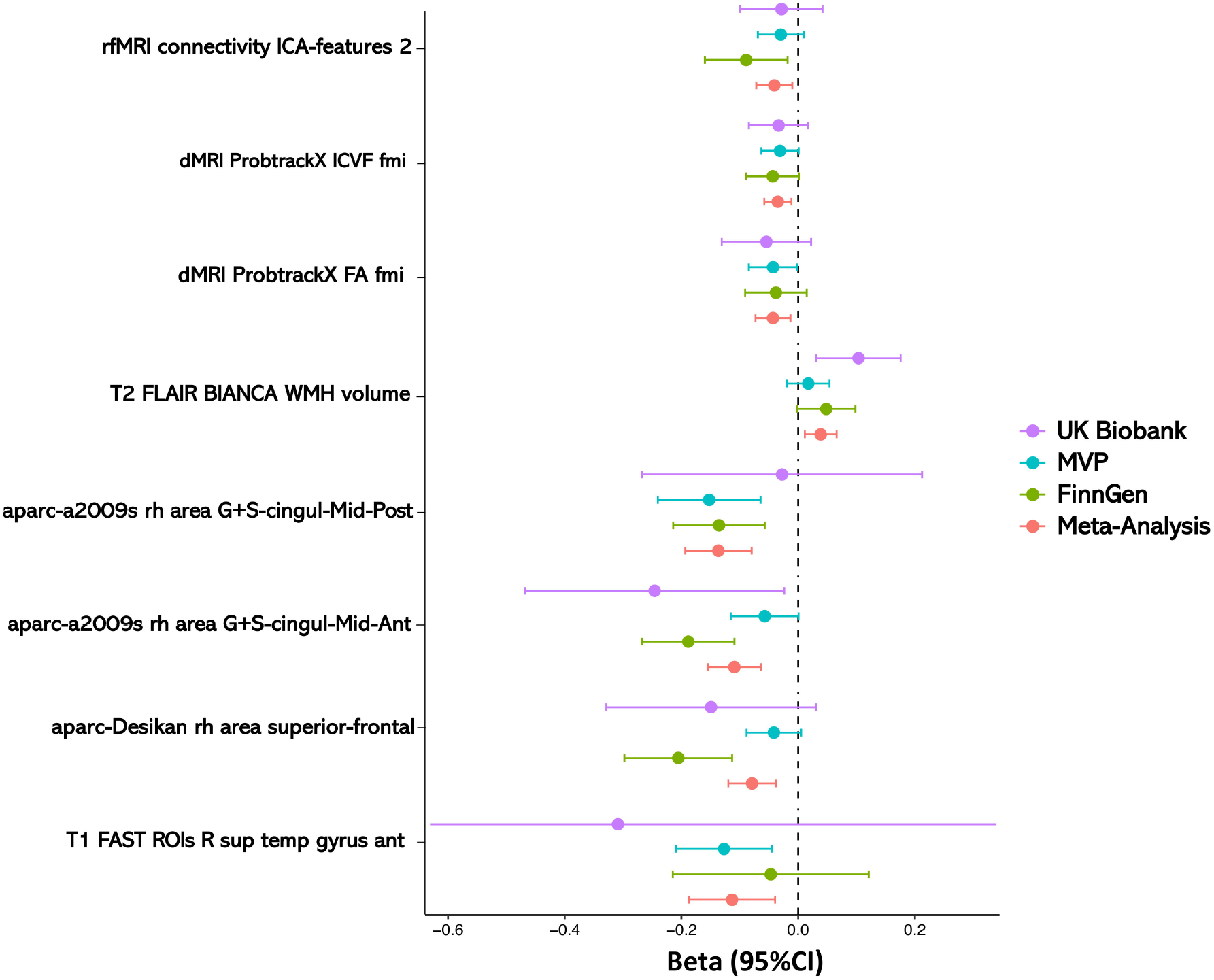
Genetically inferred effect of brain IDPs on ANX. We report the inverse-weighted variance estimates of the betas and 95% confidence intervals (CI) obtained from the two-sample Mendelian randomization (MR). T1 FAST ROIs R sup temp gyrus ant: (IDP 0043); aparc-Desikan rh area superior-frontal: (IDP 0709); aparc-a2009s rh area G+S-cingul-Mid-Ant (IDP 0952); aparc-a2009s rh area G+S-cingul-Mid-Post (IDP 0953); T2 FLAIR BIANCA WMH volume (IDP 1437); dMRI ProbtrackX FA fmi (IDP 1511); dMRI ProbtrackX ICVF fmi (IDP 1961); rfMRI connectivity ICA-features 2 (IDP 3915). Details of MR results and description of the brain IDPs are available in Supplemental Table 4.

**Figure 3.**
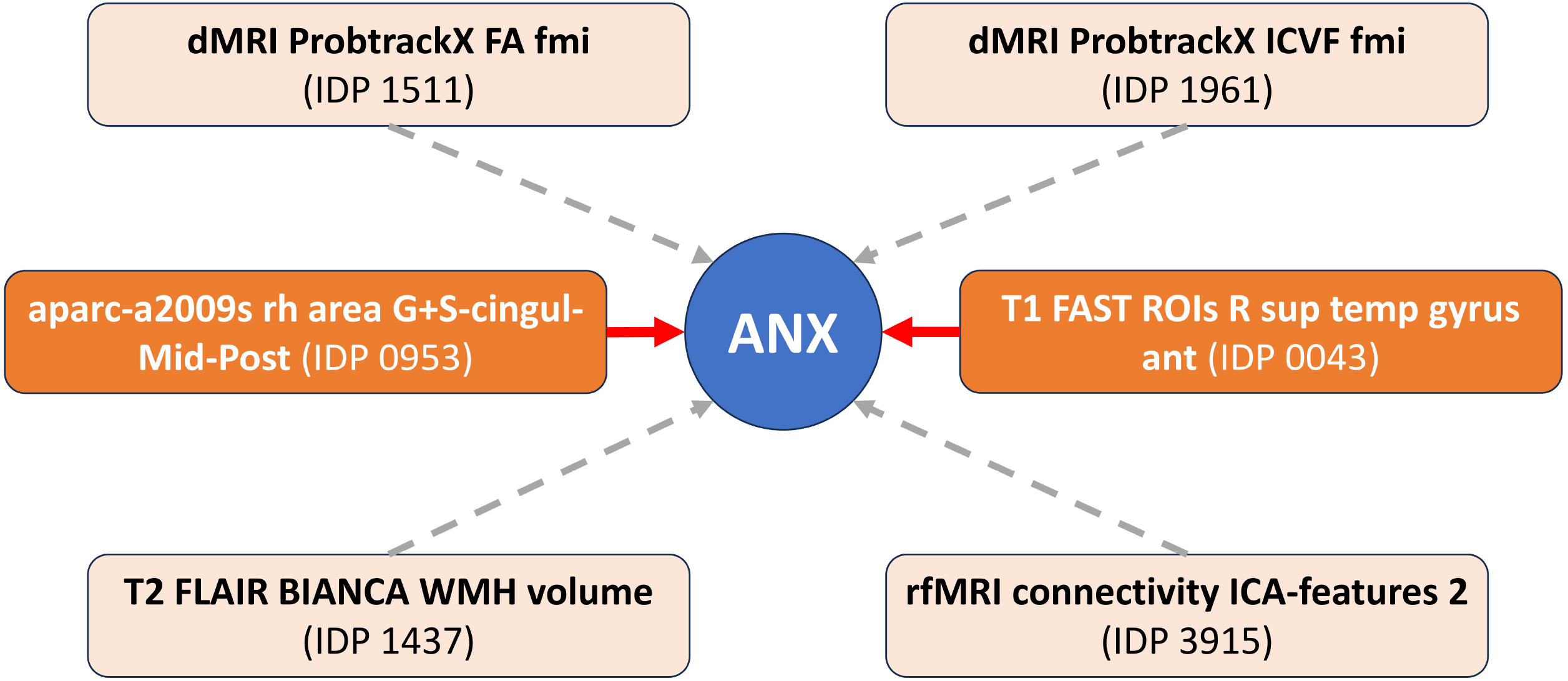
Genetically inferred direct effect of brain IDPs on ANX identified by the multivariable Mendelian randomization analysis (MVMR). IDPs with significant MVMR estimates are indicated in bright orange with red arrow. T1 FAST ROIs R sup temp gyrus ant: (IDP 0043); aparc-a2009s rh area G+S-cingul-Mid-Post (IDP 0953); T2 FLAIR BIANCA WMH volume (IDP 1437); dMRI ProbtrackX FA fmi (IDP 1511); dMRI ProbtrackX ICVF fmi (IDP 1961); rfMRI connectivity ICA-features 2 (IDP 3915). Details of MVMR results and description of the brain IDPs are available in Supplemental Table 8.

### Anxiety-Level Symptom Analysis

To explore further ANX relationships of the IPDs identified by the MVMR analysis (i.e., IDPs 0043 and 0953), we investigated possible associations with symptom-level information available from UKB (Supplemental Table 1). Additionally, because these IDPs are both related to the right hemisphere, we also investigated the corresponding IDPs on the left hemisphere (IDP 0042, the grey-matter volume in the anterior division of the left superior temporal gyrus; IDP 0879, the area of the middle-posterior part of the left cingulate gyrus and sulcus) to gain insights into the possible differences in brain lateralization. After multiple testing correction (FDR q<0.05; Supplemental Table 8), we observed that the area of the middle-posterior part of the right cingulate gyrus and sulcus (IDP 0953) was negatively associated with “tense sore or aching muscles during worst periods of anxiety” (beta=-0.13, p=8.15×10^-6^). While it did not survive multiple testing correction, the area of middle-posterior part of the left cingulate gyrus and sulcus (IDP 0879) was also nominally associated with “tense sore or aching muscles during worst periods of anxiety” (beta=-0.09, p= 0.001). In the sex-stratified analysis, we did not observe differences between females and males that survived multiple testing correction (Supplemental Table 9). Considering the IDPs identified by the MVMR analysis (i.e., IDPs 0043 and 0953), the strongest result showed opposite effect directions in the association of the anterior division of the right superior temporal gyrus (IDP 0043) with “Professional informed about anxiety” (female-beta=0.07, female-p=0.043; male-beta=-0.08, male-p-value=0.042; sex-difference p=0.004).

## DISCUSSION

Leveraging large-scale genome-wide data, the present study provided a comprehensive investigation of the pleiotropy and possible cause-effect relationships linking ANX to brain structure and function assessed across multiple MRI modalities. The genetic correlation analysis with respect to 3,446 IDPs identified multiple brain regions sharing pleiotropic effects with ANX. Notably, the most statistically significant genetic correlations were related to the left pallidum, the Broadmann Area 6, the area of the right cingulate cortex, and the area of the right precentral gyrus. These genetically informed findings are consistent with previous observational studies. For example, increased bilateral pallidum volumes were found to be associated with social anxiety disorder (SAD) in adults (44). Furthermore, negative covariation between salivary cortisol during public speaking and regional cerebral blood flow in Brodmann area 6 was reported in SAD patients (45). The anterior middle-cingulate cortex has been linked to emotion regulation (46) and the experience of negative affect and pain (47). In our genetic correlation analysis, we identified multiple IDPs related to the precentral gyrus with consistent effects across different parcellation methods (i.e., aparc-a2009s, aparc-Desikan, aparc-DKTatlas). This brain region is implicated in the planning and execution of voluntary movements and is often referred as the primary motor cortex (48, 49). With respect to ANX, the amplitude of low-frequency fluctuations (ALFF) of the right precentral gyrus was previously associated with excessive emotional status (50). This region appears to be hypoactive in anticipatory anxiety in patients with panic disorder (51). Overall, among the most strongly genetically correlated regions with ANXs, there is a pattern involving premotor/motor control.

Given that genetic correlations may be due to shared pathways or cause-effect relationships, we applied the MR approach, identifying eight brain IDPs as potentially causal for ANX. However, the effects detected for the area of the middle-anterior part of the right cingulate gyrus and sulcus and the area of the right superior-frontal gyrus showed statistically significant heterogeneity among the three cohorts investigated (i.e., UKB, MVP, and FinnGen). This suggests that cohort-specific characteristics (e.g., participants’ age, sex distribution, ANX assessment) can influence genetically inferred effects linking brain IDPs to ANX. In contrast, the other six IDPs did not show evidence of heterogeneity across datasets, indicating that their effects on ANX can be generalizable across cohorts. Because there is a certain degree of genetic correlation among these six IDPs, we performed a MVMR analysis to verify whether their effect on ANX is independent of each other.

With this approach, direct effects on ANX were observed for the area of the middle-posterior part of the right cingulate gyrus and sulcus and the grey-matter volume of the anterior division of the right superior temporal gyrus. With respect to the former, a meta-analysis of functional neuroimaging studies showed a heightened reactivity to emotional stimuli in an extended subcortical-cortical circuit, including the middle-cingulate gyrus (52). This was also linked to the response to uncertain and certain threats (anxiety and fear, respectively) (53). Evidence from rodents, monkeys, and humans (54, 55) showed that anxiety and fear may share biological mechanisms, both being processed within this subcortical-cortical circuit. Higher cerebral blood flow in the middle-cingulate gyrus was also associated with phasic-cued fear (56). Additionally, an fMRI investigation identified a substantial association between ANX severity and the ALFF in the middle-cingulate gyrus (57).

In our ANX symptom-level analysis, the area of the right posterior middle-cingulate gyrus was found to be negatively associated with tense sore or aching muscles during the worst period of anxiety. The cingulate gyrus is involved in processing pain and emotions, containing neurons with large somatic receptive fields and predominantly engaged in nociceptive activation (58). In particular, this brain region could mediate the affective-motivational component of pain processing (18). Pain-related activation and positive potential amplitudes in relation to pain and anxiety have been also reported in the right posterior middle cingulate gyrus (59, 60). Overall, these findings support the potential role of this brain region in the complex interplay among ANX, physical symptoms, and pain sensitivity.

With respect to the other brain IDPs identified by the MVMR analysis, the anterior division of the right superior temporal gyrus includes Brodmann’s 41 and 42 areas and Wernicke’s area, which are involved in sound perception and speech comprehension, respectively (61, 62). Beyond auditory processing, this brain region is also associated with emotional processing and social cognition (63). Indeed, it might be involved in social intelligence (64) and in the perception of emotions in facial stimuli (65). Additionally, functional connectivity between left and right superior temporal gyrus was reduced during the processing of ANX words compared to neutral words in affected patients (66). These findings are in line with hypotheses regarding the contribution of the superior temporal gyrus to the development of abnormal auditory perception and impaired mental organization of thought (67, 68).

Although it did not survive multiple testing correction, the strongest difference between sexes was related to the association of the anterior division of the right superior temporal gyrus with “Professional informed about anxiety”. Specifically, an affirmative reply to “Did you ever tell a professional about these problems (medical doctor, psychologist, social worker,counselor, nurse, clergy, or other helping professional)?” was associated with increased grey-matter volume in the anterior division of the right superior temporal gyrus in females, while this relationship was inverse in males. This opposite effect direction may be related to the fact that females appear to have a more positive attitude towards seeking professional assistance (69).

There are several limitations we need to acknowledge. While we identified effects that are generalizable across cohorts with different characteristics and ANX assessments, our study investigated only genetic effects generated from individuals of European descent, because of the lack of large-scale GWAS representative of other populations. Consequently, our findings may be generalizable to other ancestry groups. The genetic correlations and the effects identified between brain IDPs and ANX are relatively small, suggesting that changes across many brain regions may contribute to the disease pathogenesis. Accordingly, large sample sizes may be needed to identify the small effects linking changes in brain structure and function to ANX. Additionally, we identified only suggestive evidence of sex differences in ANX relationship with brain IDPs. This is likely due to the fact that brain regions were identified from sex-combined ANX GWAS datasets. Unfortunately, at this time female- and male-specific ANX genome-wide association statistics are not available for most of large cohorts.

In conclusion, the present study advances our understanding of the role of specific brain regions in ANX. The identified brain regions were not among the most commonly reported in the literature in the context of ANX pathogenesis. This means that their further examination could open new paths for understanding the hidden mechanisms of the disease.

## Data Availability

All data produced in the present work are contained in the manuscript

## ACKNOWLEDGEMENTS

This study was supported by grants from the National Institutes of Health (RF1 MH132337 and R33 DA047527) and One Mind. The funding sources had no role in the design of this study, its execution, analyses, interpretation of the data, and the decision to publish the results. The authors thank the participants and the investigators involved in the UK Biobank, FinnGen Project, and Million Veteran Program.

