## Supplemental Figure 1 for "Brain-Wide Mendelian Randomization Study of Anxiety Disorders and Symptoms"

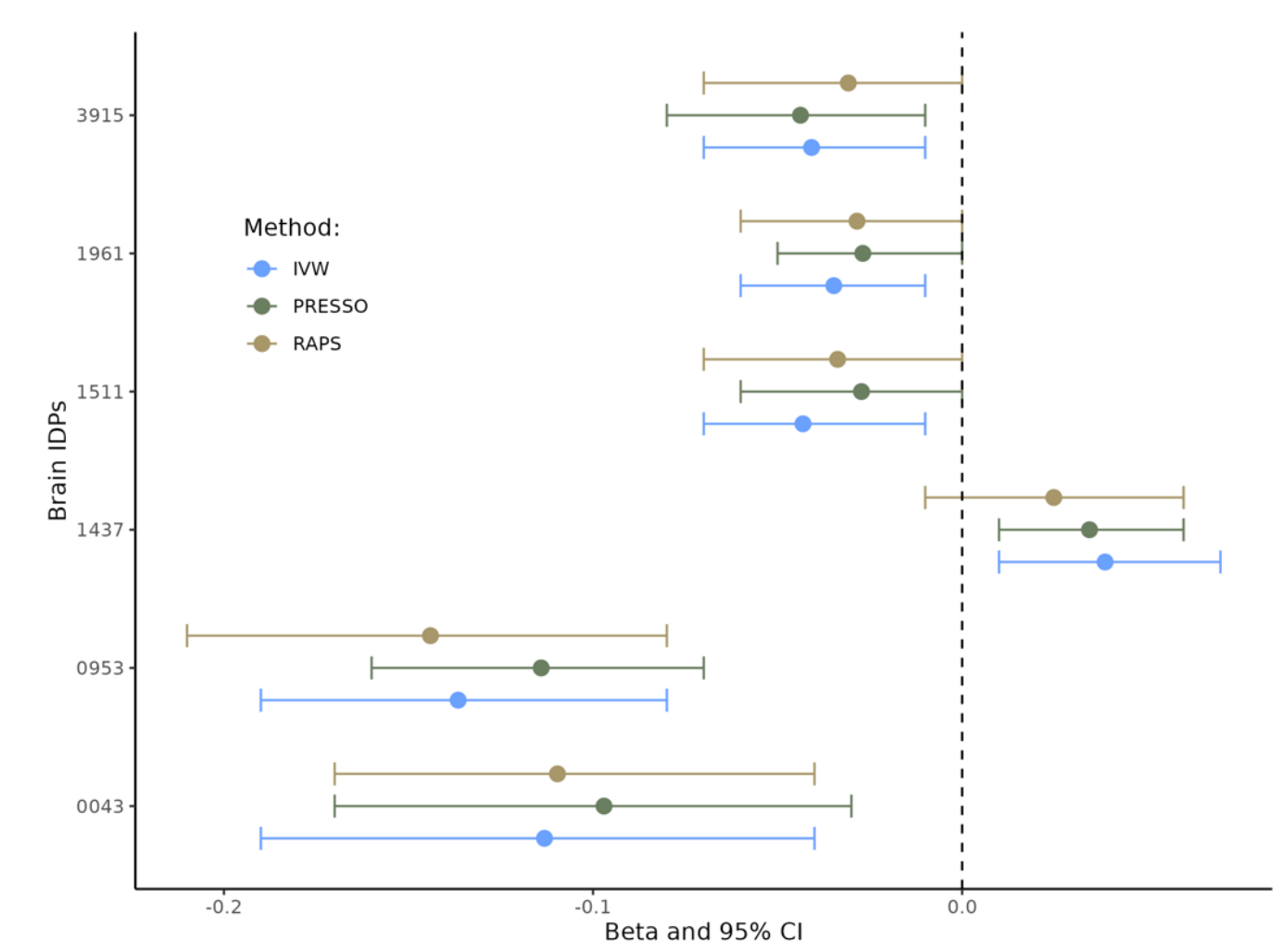

**Supplemental Figure 1. Consistency of the genetically inferred effect of brain IDPs on ANX considering different Mendelian randomization approaches.** T1 FAST ROIs R sup temp gyrus ant: (IDP 0043); aparc-a2009s rh area G+S-cingul-Mid-Post (IDP 0953); T2 FLAIR BIANCA WMH volume (IDP 1437); dMRI ProbtrackX FA fmi (IDP 1511); dMRI ProbtrackX ICVF fmi (IDP 1961); rfMRI connectivity ICA-features 2 (IDP 3915).
